# Comparing Machine Learning and Nurse Predictions for Hospital Admissions in a Multisite Emergency Care System

**DOI:** 10.1101/2025.04.07.25325126

**Authors:** Jonathan Nover, Mathew Bai, Prem Tismina, Ganesh Raut, Dhavalkumar Patel, Girish N Nadkarni, Benjamin S. Abella, Eyal Klang, Robert Freeman

## Abstract

**Background:** Emergency department (ED) crowding strains patient care and drives up costs. Early decisions on the need for patient hospital admissions can allow for better planning and potentially improve throughput and alleviate crowding. We sought to prospectively compare nurse predictions with a machine learning (ML) model for hospital admissions and to evaluate whether adding the nurse prediction to ML outputs enhances predictive performance.

**Methods:** In this prospective, observational study at six hospitals in a large mixed quaternary/community ED system (annual ED census ∼500,000), triage nurses recorded a binary admission prediction for adult patients. These predictions were compared with an ensemble ML model (XGBoost + Bio-Clinical BERT) trained on structured data (demographics, vital signs, medical history) and triage text. Nurse predictions were similarly analyzed and then integrated with the ML output to assess for improvement in predictive accuracy.

**Results:** The ensemble ML model (XGBoost + Bio-Clinical BERT) was trained on 1.8 million historical ED visits (January 2019–December 2023). It was then tested on 46,912 prospective ED visits with recorded nurse predictions (September to October 2024). In the prospective arm, nurse predictions yielded an accuracy of 81.6% (95% CI, 81.3–81.9), sensitivity of 64.8% (63.7–65.8), and specificity of 85.7% (85.3–86.0). At a 0.30 probability threshold, the ML model attained an accuracy of 85.4% (85.0–85.7) and sensitivity of 70.8% (69.8–71.7). Combining nurse predictions with the ML output did not improve accuracy beyond the model alone.

**Conclusions:** Machine learning–based predictions outperformed triage nurse estimates for hospital admissions. These findings suggest that an admission prediction system anchored by ML can perform reliably using data available at triage.

## INTRODUCTION

The effects of Emergency Department (ED) boarding have strained our healthcare system, negatively impacting patient outcomes and cost. Patient throughput in hospitals is a large and growing challenge and delays in patient progression are potentially harmful, and although health care systems are making progress most hospitals are experiencing significant operational and financial stress [1]. Such delays are often the result of patients awaiting admission decisions and bed placement in the hospital. ICU patients admitted through the Emergency Department with no delay had a lower mortality rate (12.9%) compared to ICU patients admitted with delays (17.4%) [2]. Emergency surgery patients with delays had a 53% increase in mortality, and each delayed patient had an increased length of stay of 2.6 days and increased cost of $3,335 [3]. US patients with extended hospitalizations utilize 14% to 15% of all hospital days and cost the US health Care System more than $20 billion for annuall [4].

Over the past decade artificial intelligence (AI) has entered the healthcare landscape and is being developed, studied, and used as a potential solution to improve patient throughput. More specifically, there have been studies to better understand how human and AI could accurately predict admissions in the ED, potentially providing opportunities for earlier decision making and better bed allocation planning. Alexander et al [5], published a study that showed the triage nurse correctly predicted if a patient was going to be admitted 66.9% of the time. With this low rate, they concluded that using the triage nurse to predict admissions would not help patient flow. Cameron et al [6], published a study that showed nurses correctly predicted admission 68.9% of the time and underperformed against an objective scoring tool. However the study concluded when the nurse expressed increased confidence ratings for an admission they outperformed the objective scoring tool. The authors suggested that the optimum approach seems to be a combination of the objective tool and nurse over-ruling the result when they expressed certainty.

Patel et al [7], evaluated a machine learning (ML) model that made admission predictions at multiple time points in the ED visit. This had the advantage of having more data at later timepoints and was able to achieve an Area under the Curve (AUC) of 0.81-0.88. Hong et al [8], compared prediction accuracy using triage only data against using available medical history in the electronic health record (EHR). They found that adding historical data boosted their AUC from 0.88 to 0.93, when compared to using triage-only data. A recent publication in Mayo Clinic Proceedings Digital Health, by Williams et al [9], reviewed the literature on this acute subject. The study explains that including observation unit or short stay admissions to the machine learning model can lead to significant improvements, boosting an AUROC of 0.86 to 0.93. With these improved rates of admission prediction when using AI, these predictions may finally be accurate enough to be useful in improving patient throughput for patients that are admitted.

Our aim was to prospectively compare a machine learning approach and nurse predictions for hospital admissions across a large, diverse healthcare system to potentially demonstrate a prediction model with high enough accuracy to be used to augment bed planning.

## METHODS

### Study and Setting

This was a prospective, observational study conducted within the Emegency Departments of the Mount Sinai Health System in New York, encompassing six hospitals. The 6 EDs have a combined annual volume of approximately 500,000 patients annually, and span from quaternary academic medical center, trauma center, to the community hospital setting. Three hospitals are 911 STEMI receiving sites, two sites Thrombectomy Capable Stroke and one site Comprehensive Stroke Center. All sites are American College of Emergency Physician (ACEP) Geriatric certified, silver or bronze, three sites achieved the Emergency Nurses Association (ENA) Lantern Award, and two sites are American Nursing Credentialing Center (ANCC) Magnet designated.

The study evaluated hospital admission predictions made at ED triage by nurses and a machine learning (ML) model. Data collection spanned two months, from September to October 2024. The study was approved by the Institutional Review Board (IRB) at the Icahn School of Medicine at Mount Sinai. (IRB-18-00573-MODCR001)

Patients were included if they presented to the ED during the study period and underwent standard triage. Transfer to another facility directly from the ED was considered as hospital admission. **Figure 1** outlines the overall study design, from the initial dataset and model training to the prospective collection of nurse predictions and final performance evaluation.

**Figure 1:**
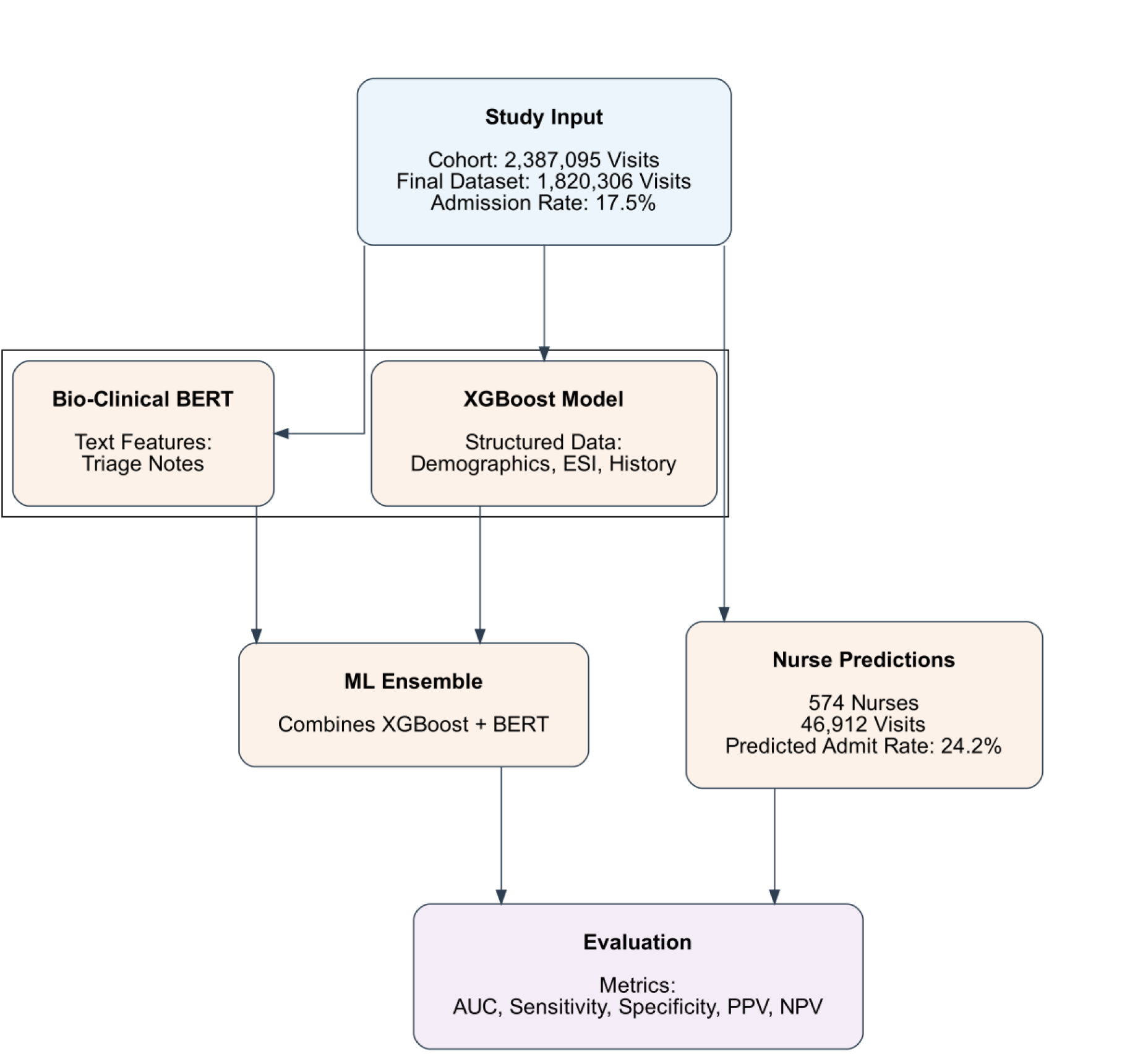
Study Workflow for Admission Prediction. The figure illustrates how a large cohort of emergency department (ED) visits (final dataset of 1,820,306 visits) was divided into structured (demographics, ESI, history) and unstructured (triage text) inputs. These were processed by XGBoost and Bio-Clinical BERT models, respectively, and combined into an ensemble. Prospective nurse predictions from 46,912 ED visits were separately recorded and then compared with the ensemble’s output during the evaluation phase.

### Nurse-Based Admission Predictions

As part of this study, ED triage nurses were required to document a “Yes” or “No” decision for each patient at the end of the triage process. This decision step was incorporated into the electronic triage system workflow to standardize prediction collection across facilities. The nurses whom performed the triage during the study period had at least 1 year of ED experience and ranged as high as 30 years of ED experience and completed the Emergency Severity Index (ESI) course and training which included at least 12 hours of triage preceptorship. Additionally, all triage RNs were trained about the study by reviewing a Nursing Practice Alert (**Figure 2**). All documented decisions were later extracted from the triage system for comparison with ML predictions. A disposition of ED Observation was not counted as an admission. Our hospital system observation units are staffed and managed by the ED, within the physical footprint of the ED.

**Figure 2:**
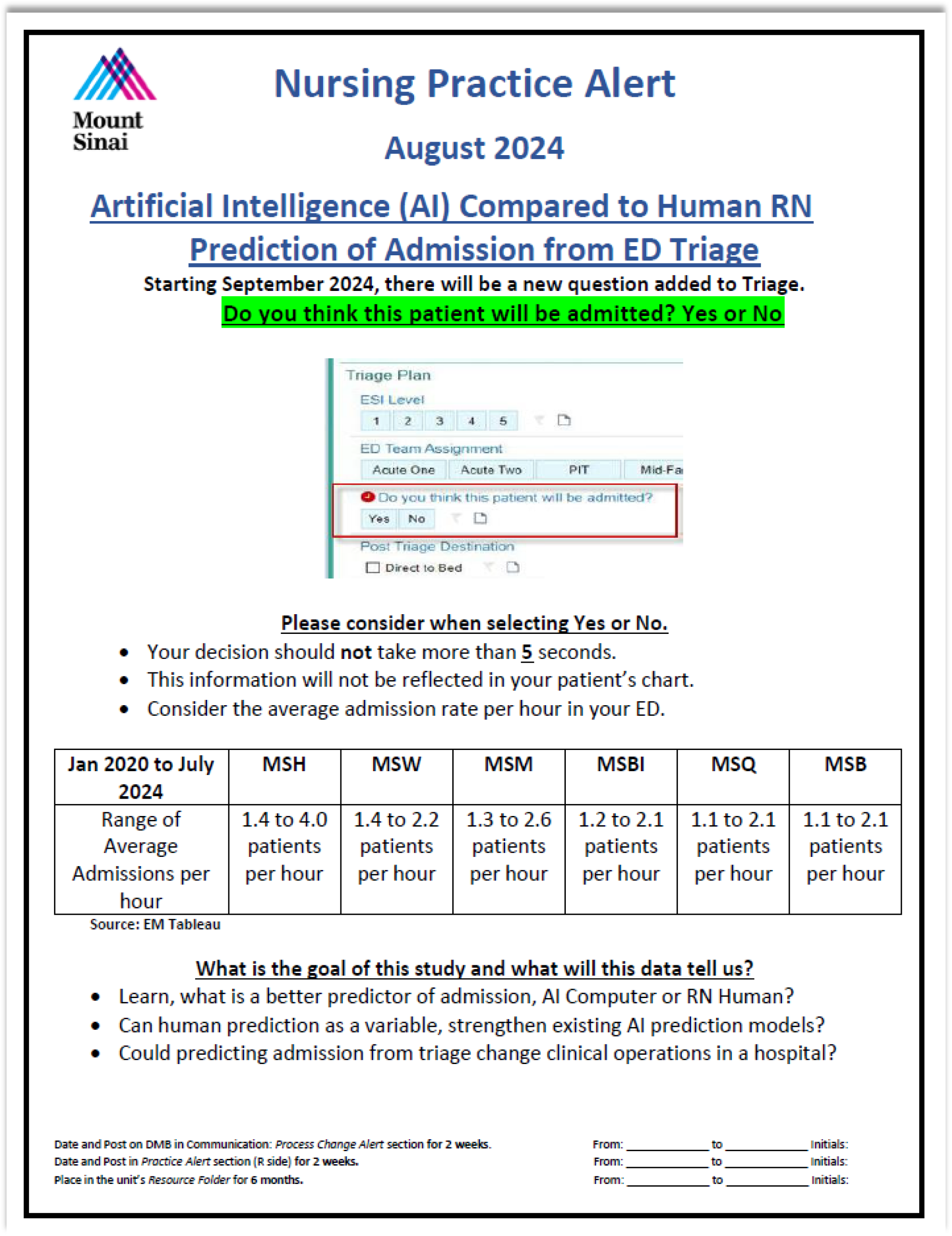
Triage RNs education about the study using a Nursing Practice Alert.

### Machine Learning Model

The ML model was designed to predict hospital admission at triage based on structured and unstructured patient data. Structured data inputs included demographics (age, race, sex), vital signs, Emergency Severity Index (ESI) score, history of ICD-10-CM codes (grouped using Clinical Classifications Software [CCS] **Clinical Classification Software (CCS) for ICD-10-CM Fact Sheet** [10]), counts of prior ED visits, and hospitalizations, as well as the chief complaint. Triage notes were used as unstructured text data.

The model architecture comprised an average ensemble of XGBoost algorithms for structured data and a BERT-based natural language processing model for text data, following a previously published approach by Glicksberg BS *et al.*[11]. Feature engineering included transforming categorical variables, and computing counts of prior ED visits / counts of prior hospitalizations. Missing data was handled through XGBoost functionality for structured data, and filling of empty strings for textual data. Model training was conducted using (PyTorch and scikit-learn in Python 3.9).

### Model Development and Validation

Model training data comprised ED visit records from January 2019 to December 2023. Validation was conducted on data from January to August 2024, and testing was performed on data from September and October 2024 to allow direct comparison with nurse predictions. Cross-validation was performed during model training to select hyperparameters. Data preprocessing and hyperparameter tuning were conducted using grid search with a five-fold cross-validation approach.

All computations were performed on Mount Sinai Health System clusters with NVIDIA H100 GPUs [12], each with 80 GB of memory, running Python 3.9. The final model was selected based on AUC performance in the validation set.

An overview of our end-to-end workflow, including data ingestion, ensemble modeling (XGBoost for structured data and Bio-Clinical BERT for text data), and the integration of prospective nurse predictions, is provided in **Figure S1 in the supplementary file**.

### Enhanced Predictive Accuracy through Combined Human-AI Analysis

To evaluate the predictive accuracy of combined nurse and machine learning (ML) model assessments, we conducted a comparative analysis focusing on positive predictive value (PPV) for patient admissions. First, we generated a confusion matrix to distinguish cases where both the nurse and the ML model flagged patients for admission versus cases where only one or neither predicted admission. We then assessed the subset of cases with concordant (both predicted admission) and discordant (one predicted, one did not) outcomes, with a particular focus on the PPV for concordant cases. By comparing PPVs across nurse-only, ML-only, and combined predictions, we aimed to determine if cases flagged by both sources correspond to a higher probability of actual admissions, thus supporting the hypothesis that human-AI synergy could yield a more reliable admission prediction. This analysis serves to quantify the benefit of dual-assessment approaches in enhancing admission prediction accuracy.

### Statistical Analysis

Model performance metrics included area under the receiver operating characteristic curve (AUC), accuracy, F1 score, sensitivity, specificity, positive predictive value (PPV), and negative predictive value (NPV), for different probability cut-off values, and Youden’s index. Each reported metric had a 95% confidence interval (CI) estimation using 1,000 bootstrapped samples. Nurse predictions, being binary (admit/discharge), were evaluated with accuracy, F1 score, sensitivity, specificity, PPV, and NPV.

Statistical comparisons of performance between the ML model and nurse predictions were conducted using the McNemar’s test for paired binary metrics (sensitivity, specificity, PPV, and NPV).

## RESULTS

### Data Characteristics

#### Overall Cohort

Across the Mount Sinai Health System emergency departments from January 2019 to October 2024, the initial cohort comprised 2,387,095 patient visits, representing 817,644 unique patients. After excluding visits with missing age data (n=283,452), the cohort was reduced to 2,103,643 visits. Finally, removing visits with incomplete text documentation yielded a final dataset of 1,820,306 visits. The overall admission rate for the entire cohort was 17.5%.

**Figure 3** illustrates the integrated cohort design and the nurse prediction workflow for ED admission assessment, detailing the data inclusion and exclusion criteria for both the overall cohort and the nurse prediction set.

**Figure 3:**
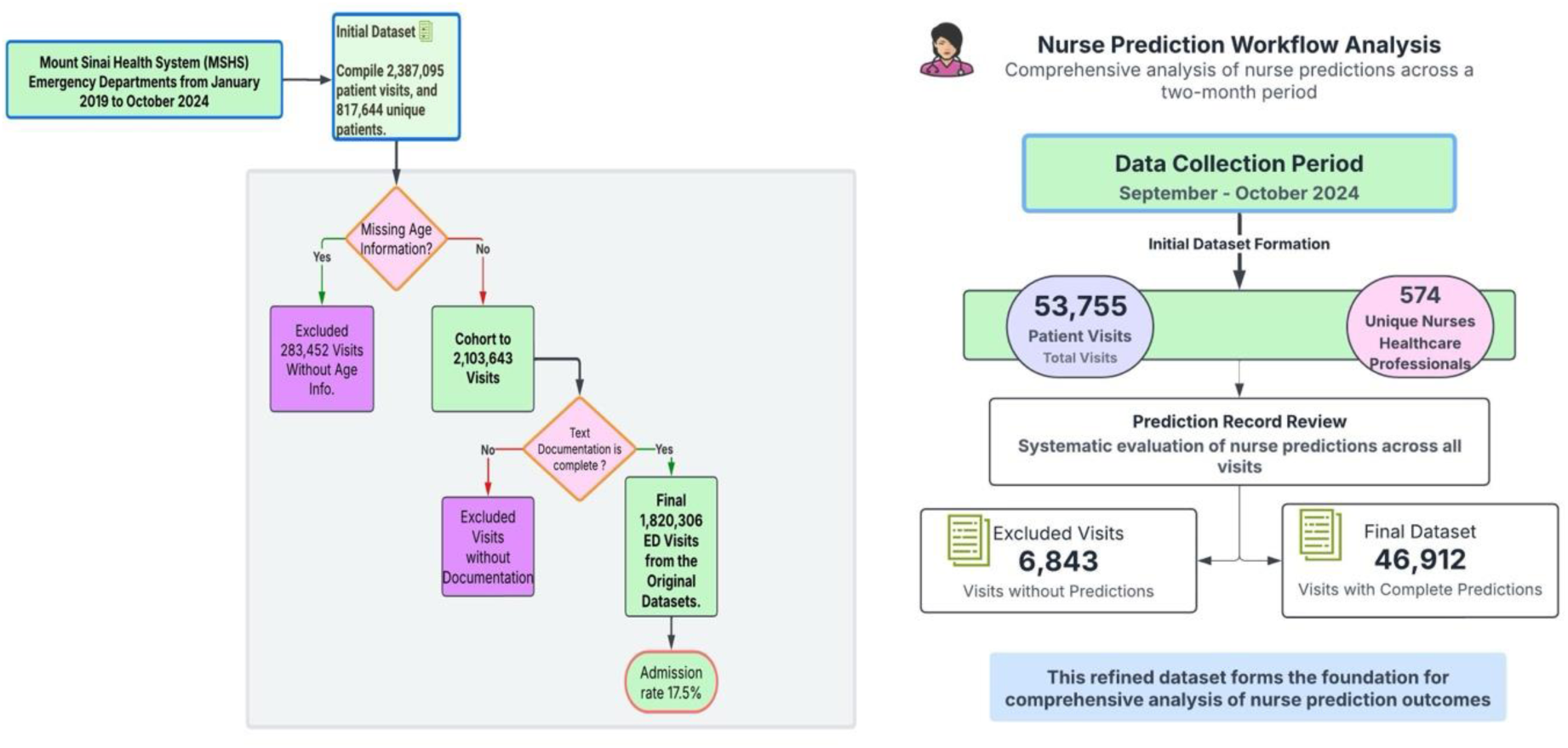
Cohort Design and Nurse Prediction Workflow for ED Admission Assessment

#### Nurse Prediction Set

For the nurse prediction component, data were collected over a two-month period from September to October 2024. This subset initially included 53,755 patient visits, with predictions recorded by 574 unique nurses. Of these visits, 46,912 included recorded nurse predictions, forming the final nurse prediction set. Nurses predicted admissions at a rate of 24.2%, which was higher than the actual observed admission rate.

### Patient Characteristics by Admission Status

Key patient characteristics differed significantly between discharged and admitted patients (**Table 1**). Admitted patients were notably older, with a median age of 63.0 years compared to 38.7 years for discharged patients. A higher proportion of male patients were admitted, and there were distinct racial distributions, with White and Asian patients more commonly admitted.

**Table 1.** demographic and clinical characteristics of patients by admission status. Values represent median (IQR) for continuous variables and counts (%) for categorical variables.

| Variable | Discharged<br>(n=1,501,012, 82.5%) | Admitted<br>(n=319,294, 17.5%) | P-value |
| --- | --- | --- | --- |
| Age (years) | 38.7 (25.8 - 57.7) | 63.0 (44.8 - 76.4) | <0.001 |
| Sex | Female: 776,479 (51.7%); Male: 724,363 (48.3%); Unknown/Indeterminate: 170 (0.0%) | Male: 161,612 (50.6%); Female: 157,660 (49.4%); Unknown/Indeterminate: 22 (0.0%) | <0.001 |
| Race | Other: 700,014 (46.6%); Black: 423,154 (28.2%); White: 302,638 (20.2%); Asian: 75,206 (5.0%) | Other: 125,617 (39.3%); White: 91,025 (28.5%); Black: 84,917 (26.6%); Asian: 17,735 (5.6%) | <0.001 |
| Emergency Severity Index (ESI) | 3.0 (3.0 - 4.0) | 3.0 (2.0 - 3.0) | <0.001 |
| Systolic Blood Pressure (mmHg) | 130.0 (117.0 - 145.0) | 133.0 (117.0 - 152.0) | <0.001 |
| Pulse (beats/min) | 86.0 (75.0 - 98.0) | 89.0 (76.0 - 104.0) | <0.001 |
| Pain Score (0 - 10) | 5.0 (0.0 - 8.0) | 4.0 (0.0 - 8.0) | <0.001 |

#### Chief Complaints by Admission Status

Chief complaints showed distinct patterns by admission status (**Supplementary Table S1**). Among admitted patients, “Shortness of Breath” (9.3%) and “Abdominal Pain” (9.3%) were the most common specific complaints, while “Unspecified” complaints accounted for 9.9%. In contrast, “Abdominal Pain” (7.9%) and “Chest Pain” (4.4%) were more frequent among discharged patients, alongside a higher proportion of “Unspecified” cases (10.9%).

#### Past Medical History by Admission Status

The analysis of past medical histories, categorized using Clinical Classification Software (CCS), highlighted notable patterns between discharged and admitted patients (**Supplementary Table S2**). Among discharged patients, the most frequent histories involved “Factors Influencing Health Status” (FAC) and “Symptoms, Signs, and Abnormal Findings” (SYM), accounting for 13.7% and 13.6% of cases, respectively. For admitted patients, the most common categories included “Symptoms” (SYM) at 13.4% and “Diseases of the Circulatory System” (CIR) at 12.5%, indicating common health conditions linked to higher admission rates.

#### Emergency Severity Index (ESI) and Admission Rates

Admission rates were highest in high-acuity patients, with 64.8% for ESI 1 and 38.6% for ESI 2. In contrast, lower-acuity levels showed sharply reduced admission rates, with 16.0% for ESI 3, 1.9% for ESI 4, and 0.5% for ESI 5. These results highlight a clear trend of increased admission likelihood with higher ESI severity (**Table 2**, **Figure 4**).

**Table 2:** distribution of true admission rates and nurse admission predictions across Emergency Severity Index (ESI) levels.

| ESI Level | True Admissions / Total Patients (%) | Nurse Predicted Admissions (%) |
| --- | --- | --- |
| 1 | 9,126 / 14,088 (64.8%) | 196/329 (59.6%) |
| 2 | 140,293 / 363,734 (38.6%) | 4,004/9,912 (40.4%) |
| 3 | 157,545 / 985,946 (16.0%) | 4,400/26,512 (16.6%) |
| 4 | 6,906 / 367,072 (1.9%) | 169/8,332 (2.0%) |
| 5 | 241 / 47,136 (0.5%) | 4/616 (0.6%) |

**Figure 4:**
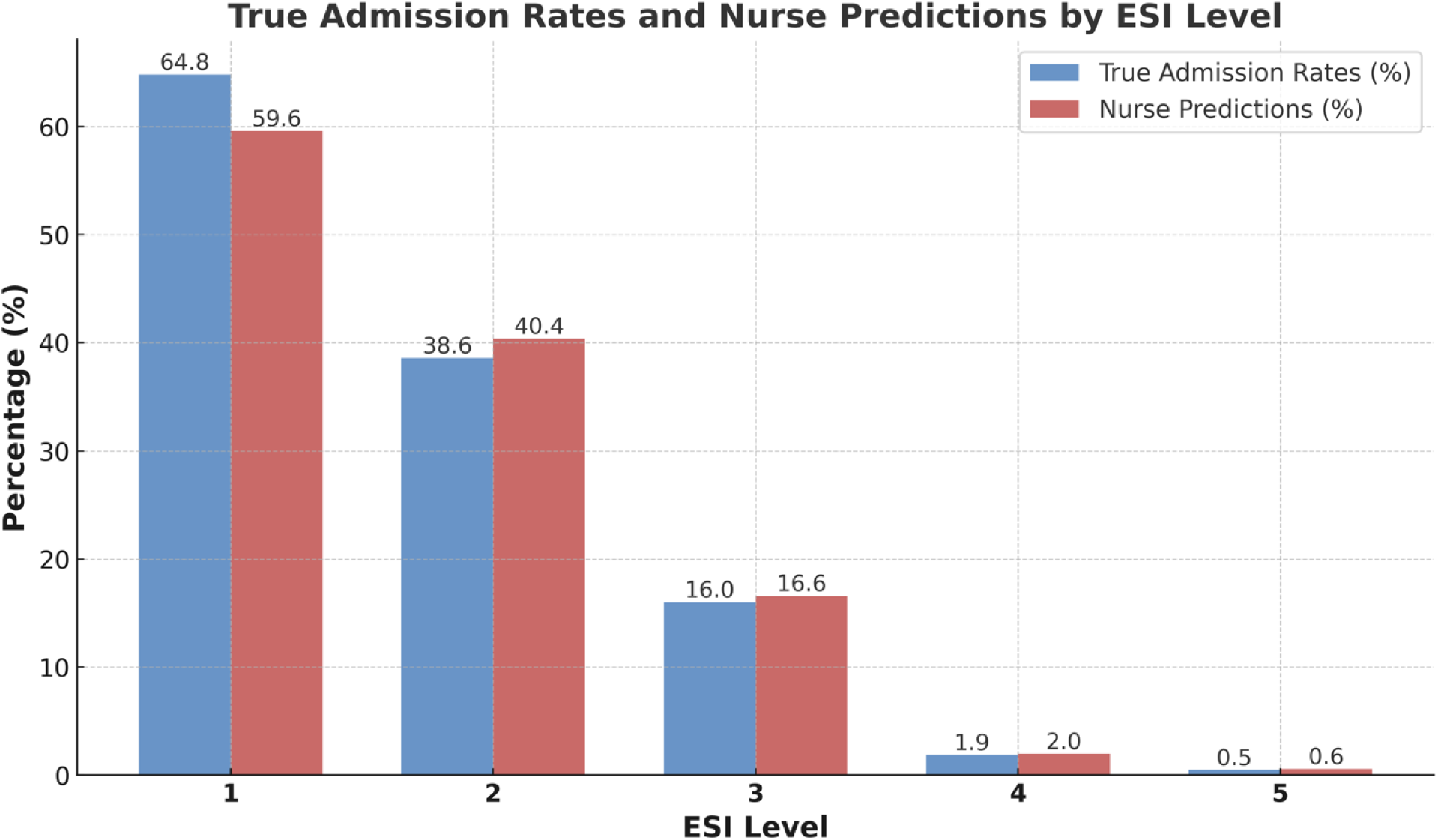
True admission rates and nurse predictions by ESI Level.

#### Nurse Predictions by Emergency Severity Index (ESI)

Nurse predictions varied across ESI levels, reflecting a pattern similar to actual admission rates. For ESI 1, nurses predicted admissions for 59.6% of patients, and for ESI 2, predictions were 40.4%. Predictions dropped in lower acuity levels, with rates of 16.6% for ESI 3, 2.0% for ESI 4, and 0.6% for ESI 5 (**Table 2**, **Figure 4**).

#### Machine Learning Model

Full details of the machine learning hyperparameter tuning are provided in **Supplementary Tables S3–S5**. For XGBoost, we evaluated combinations of n_estimators, learning_rate, and max_depth, with the highest AUC (0.871) achieved at n_estimators=1000, learning_rate=0.1, and max_depth=6. Single-feature analysis (**Supplementary Figure S2**) revealed that medical history (CCS set), age, and ESI were the strongest individual predictors. For Bio-Clinical BERT, we explored various learning rates, batch sizes, and weight decay settings; a learning rate of 1×10^−5^, batch size of 8, and weight decay of 0.01 yielded the best AUC (0.840). However, in practice, we adopted a higher batch size (256) for computational efficiency, resulting in a near-optimal AUC of 0.838.

#### XGBoost Model Performance at Different Thresholds

The final XGBoost model achieved an AUC of 0.875. **Supplementary Table S6** presents the XGBoost model’s predictive performance metrics—accuracy, F1 score, sensitivity, specificity, negative predictive value (NPV), and positive predictive value (PPV)—are shown across various probability thresholds for admission.

#### NLP Model Performance at Different Thresholds

The NLP model achieved an AUC of 0.862. **Supplementary Table S7** presents the performance metrics of the Natural Language Processing (NLP) model across various probability thresholds for predicting admission. The metrics include accuracy, F1 score, sensitivity, specificity, negative predictive value (NPV), and positive predictive value (PPV). Performance trends align with expected trade-offs between sensitivity and specificity at different thresholds.

#### Ensemble Model (XGBoost + Bio-Clinical BERT) Performance

The ensemble model, combining XGBoost and Bio-Clinical BERT, demonstrated superior predictive performance across all thresholds. **Table 3** presents the ensemble’s performance metrics, including accuracy, F1 score, sensitivity, specificity, NPV, and PPV. The model achieved an AUC of 0.894, showcasing robust discrimination between admitted and non-admitted patients.

**Table 3:** Performance metrics of the ensemble model at different probability thresholds.

| Threshold | Accuracy | F1 Score | Sensitivity | Specificity | NPV | PPV |
| --- | --- | --- | --- | --- | --- | --- |
| 0.10 | 64.7% (64.3% - 65.2%) | 0.51 (0.50 - 0.52) | 94.3% (93.9% - 94.8%) | 57.6% (57.1% - 58.1%) | 97.7% (97.5% - 97.9%) | 34.9% (34.4% - 35.5%) |
| 0.19 (Youden’s index) | 79.4% (79.0% - 79.7%) | 0.61 (0.61 - 0.62) | 84.4% (83.7% - 85.2%) | 78.1% (77.7% - 78.6%) | 95.4% (95.2% - 95.6%) | 48.2% (47.4% - 49.0%) |
| 0.30 | 85.4% (85.0% - 85.7%) | 0.65 (0.64 - 0.66) | 70.8% (69.8% - 71.7%) | 88.9% (88.6% - 89.2%) | 92.6% (92.4% - 92.9%) | 60.6% (59.6% - 61.5%) |
| 0.50 | 86.7% (86.5% - 87.1%) | 0.57 (0.56 - 0.58) | 45.3% (44.3% - 46.3%) | 96.7% (96.6% - 96.9%) | 88.0% (87.7% - 88.3%) | 77.1% (75.9% - 78.2%) |
| 0.70 | 84.1% (83.8% - 84.5%) | 0.34 (0.33 - 0.35) | 21.2% (20.4% - 22.1%) | 99.3% (99.2% - 99.4%) | 83.9% (83.6% - 84.3%) | 88.4% (87.0% - 89.7%) |
| 0.90 | 81.3% (80.9% - 81.6%) | 0.07 (0.07 - 0.08) | 3.9% (3.5% - 4.3%) | 100.0% (99.9% - 100.0%) | 81.2% (80.8% - 81.5%) | 96.2% (94.2% - 97.9%) |

**Table 4** evaluates the model’s performance across probability thresholds for predicting patient admissions. Sensitivity, positive predictive value (PPV), and confusion matrix elements (TP, FP, TN, FN) are provided. The percentage of the total population identified represents the proportion of admitted patients detected relative to the entire dataset.

**Table 4:** Performance metrics across probability thresholds for the ensemble model.

| Threshold | Sensitivity (%) | PPV (%) | TP | FP | TN | FN | % of Total Population Identified |
| --- | --- | --- | --- | --- | --- | --- | --- |
| 0.1 | 94.3 | 34.9 | 8607 | 16026 | 21763 | 516 | 18.3 |
| 0.2 | 83.3 | 49.5 | 7596 | 7742 | 30047 | 1527 | 16.2 |
| 0.3 | 70.8 | 60.6 | 6455 | 4195 | 33594 | 2668 | 13.8 |
| 0.4 | 57.9 | 69.3 | 5285 | 2336 | 35453 | 3838 | 11.3 |
| 0.5 | 45.3 | 77.1 | 4137 | 1230 | 36559 | 4986 | 8.8 |
| 0.6 | 32.4 | 83.2 | 2955 | 595 | 37194 | 6168 | 6.3 |
| 0.7 | 21.2 | 88.4 | 1938 | 255 | 37534 | 7185 | 4.1 |
| 0.8 | 10.9 | 92.8 | 996 | 77 | 37712 | 8127 | 2.1 |
| 0.9 | 3.9 | 96.2 | 354 | 14 | 37775 | 8769 | 0.8 |

#### Nursing Predictions Performance

Nursing predictions achieved an accuracy of 81.6%, with a sensitivity of 64.8% and specificity of 85.7% (**Table 5**).

**Table 5:** performance metrics for nursing predictions on admission outcomes.

| Metric | Value |
| --- | --- |
| Accuracy | 81.6% (81.3% - 81.9%) |
| F1 Score | 0.58 (0.57 - 0.59) |
| Sensitivity | 64.8% (63.7% - 65.8%) |
| Specificity | 85.7% (85.3% - 86.0%) |
| NPV | 91.0% (90.7% - 91.3%) |
| PPV | 52.2% (51.3% - 53.1%) |

#### Comparison of Ensemble Model and Nursing Predictions

**Figure 5** illustrates the ROC curves for the ensemble model, Bio-Clinical BERT (NLP), and XGBoost, compared against the single-point performance of nurses. The ensemble model achieved the highest AUC (0.894), followed by NLP (0.862) and XGBoost (0.875). The nurse predictions align with a sensitivity of 64.8% and specificity of 85.7%, visualized as a distinct point on the curve.

**Figure 5:**
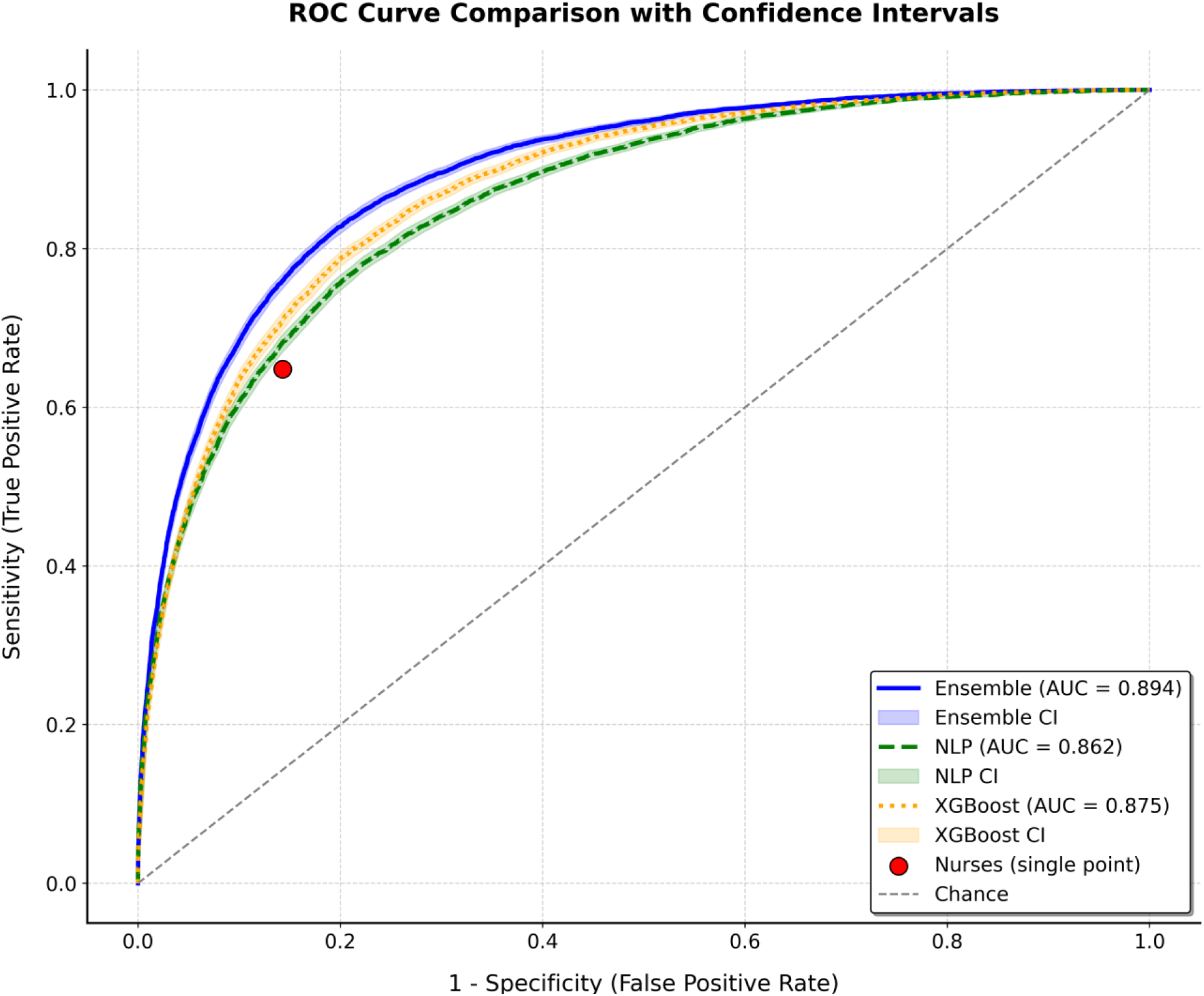
ROC curves comparing the ensemble model (AUC = 0.894), NLP (AUC = 0.862), and XGBoost (AUC = 0.875) with the performance point of nurses.

The ensemble model outperformed nursing predictions in several key metrics. At a threshold of 0.30, the ensemble achieved an accuracy of 85.4%, compared to the nurses’ 81.6%. Sensitivity at this threshold was also higher for the ensemble (70.8% vs. 64.8%), as was PPV (60.6% vs. 52.2%). These results suggest that the ensemble model provides better overall predictive performance, particularly in its ability to identify true admissions (sensitivity) and make accurate positive predictions (PPV).

#### Integration of Nurse Predictions into the Ensemble Model

We tested an ensemble that combined the machine learning model output and the nurses’ binary predictions. For the ensemble, we used a threshold of 0.5 for the model’s predicted probability and incorporated the nurse prediction as a binary vote. If over half of the combined votes supported admission, the ensemble predicted admission.

The ensemble incorporating nurses’ predictions achieved an accuracy of 86.2%. In contrast, the machine learning model alone (at a 0.5 probability threshold) achieved an accuracy of 86.7%. Thus, adding nurses’ predictions did not improve overall accuracy. This result suggests that nurse-based predictions did not provide incremental value when integrated with the model’s output.

## DISCUSSION

Rather than seek an AI alternative to clinical judgement, we sought to augment the human factor with AI. When comparing the human and AI prediction, the AI models outperformed the human prediction by a small margin. Additionally, although the human-in-the-loop (HITL) concept did not increase the accuracy of predicting admissions beyond what is currently possible with only the nurse or the AI model, when we added the human prediction to the AI model, we found that the results remained fairly constant. Williams et al [9] meta-analysis expressed an overall accuracy of triage nurse prediction from triage between 73% to 80.6%. Compared to the Williamson survey, this study displayed an accuracy of triage nurse prediction of 81.6%, hence outperforming the results expressed in the literature. Our ensemble model achieved an accuracy of 86.7%. Compared to groups that have researched admission prediction models, our model achieved similar results by Patel et al, [7]. Of note, Hong et al [8], had a model that was able to achieve a higher AUC of up to 0.93 by leveraging complex EMR data to gather medical history of each patient. That study used almost a thousand features in the model such as prior ECG and lab data. The disadvantages of this could be anticipated to be increased compute requirements, increase in processing time and risk of over-fitting.

Our model uses information that is gathered during routine triage in the ED as well as past visits records. The admission prediction information could be used to initiate steps in the admission process hours earlier than would otherwise be possible since we are not waiting for a provider to finish evaluating the cases; theoretically creating a reservation list to allow for bed planning in advance. For this to be operationally advantageous, the prediction model would need to have a high PPV or else we would risk holding beds for patients that are discharged from the ED, while other admitted boarding patients are still waiting in the Emergency Department.

There have been groups that attempted to operationalize admission prediction models to improve patient flow. Peck et al [13], used a prediction model that had an AUC 0.887, which approaches our model’s performance. They found that, on average, they were able to predict peak admissions about 3.5 hours before it happened. This could give the inpatient team advanced notice to upstaff if they are able to anticipate peak times accurately.

Future work will include modeling novel workflows using the AI and HITL predictive admission information. If we can demonstrate a reduction in ED Length of Stay (LOS) and boarding time as a result, this may garner support for quality improvement piloting in the live clinical environment. Additionally, we may also evaluate if this tool is useful for helping the inpatient teams anticipate their needs and start increasing staffing prior to a predicted admission peak, which would also have an impact on ED crowding and patient safety [14].

Our study limitations include use of a single health system, which limits broad generalization. The prospective data window spanned only two months, so longer-term trends may differ. We only gathered binary nurse predictions, and we did not track how this study might have changed clinical behavior. The model relied on structured triage data and free-text notes; it omitted labs or imaging results available later in the visit. Real-time, fully integrated performance was not tested. Finally, our approach focused on predictive accuracy but did not assess bed use, staffing, or cost outcomes.

In conclusion, AI outperformed Nursing prediction. When nursing prediction was added as a variable to the AI model, the results remained fairly constant. This leaves one to further ponder if an AI HITL model is the right path to admission prediction, if a different mode of incorporating nursing predictions is better, or if AI models alone are sufficient. Increasing the PPV of any prediction models is crucial to having an operational impact on the emergency department. However, if admission prediction models are unable to achieve a level of PPV to be advantageous to bed planning, it may still allow the inpatient team to anticipate an admission peak and initiate efforts to upstaff several hours in advance.

## Data Availability

All data produced in the present study are available upon reasonable request to the authors

https://icahn.mssm.edu/about/departments-offices/ai-human-health

## Acknowledgements

Romona Tulloch, Ashley Caceres, Jill Frick, Anthony Duncan, Ledjan Halollari, Louis Calderon, Shari Weisberg

---

**Supplementary Table S1:**
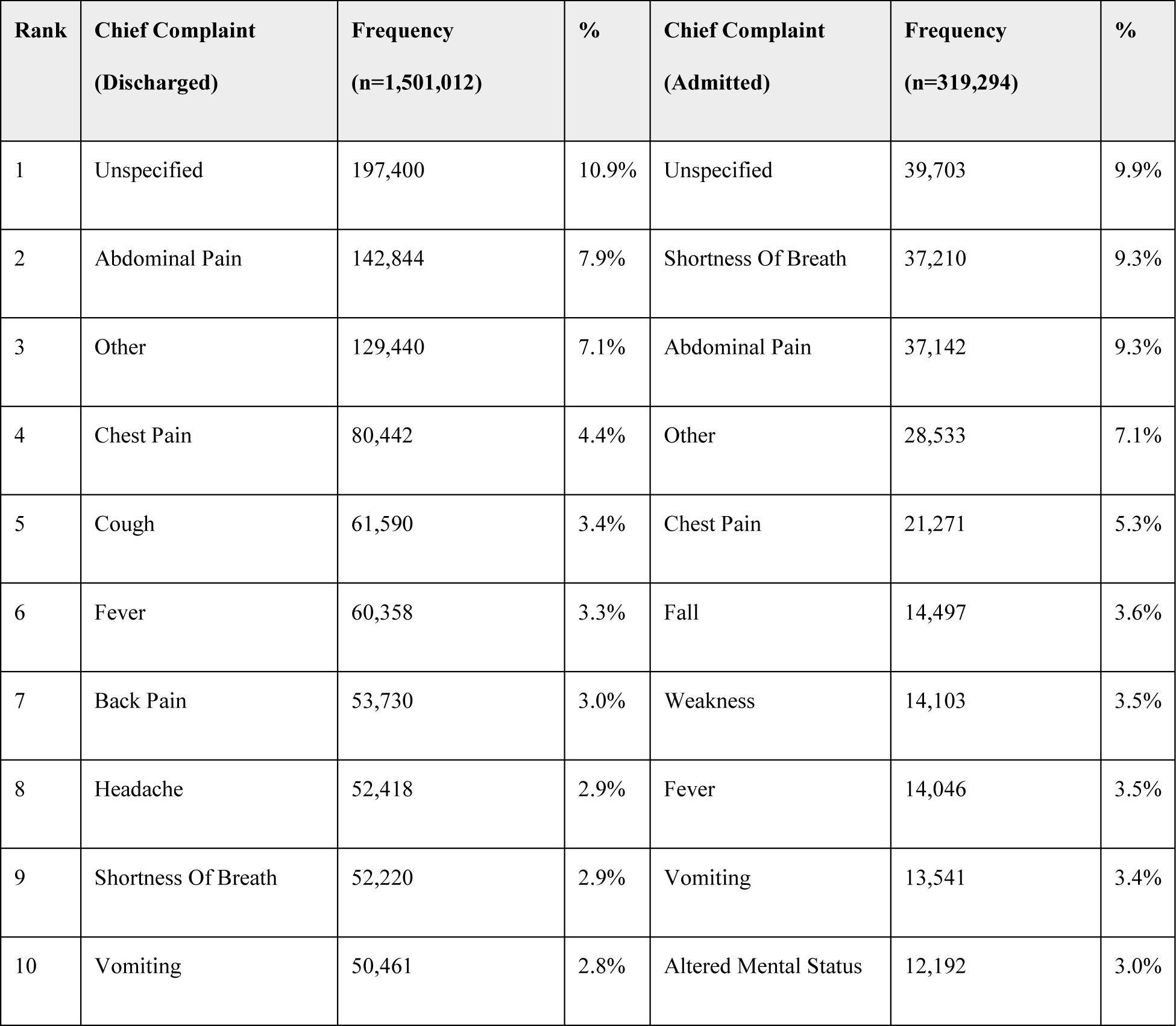
the ten most common chief complaints among patients by admission status, ranked by frequency.

| Rank | Chief Complaint<br>(Discharged) | Frequency<br>(n=1,501,012) | % | Chief Complaint<br>(Admitted) | Frequency<br>(n=319,294) | % |
| --- | --- | --- | --- | --- | --- | --- |
| 1 | Unspecified | 197,400 | 10.9% | Unspecified | 39,703 | 9.9% |
| 2 | Abdominal Pain | 142,844 | 7.9% | Shortness Of Breath | 37,210 | 9.3% |
| 3 | Other | 129,440 | 7.1% | Abdominal Pain | 37,142 | 9.3% |
| 4 | Chest Pain | 80,442 | 4.4% | Other | 28,533 | 7.1% |
| 5 | Cough | 61,590 | 3.4% | Chest Pain | 21,271 | 5.3% |
| 6 | Fever | 60,358 | 3.3% | Fall | 14,497 | 3.6% |
| 7 | Back Pain | 53,730 | 3.0% | Weakness | 14,103 | 3.5% |
| 8 | Headache | 52,418 | 2.9% | Fever | 14,046 | 3.5% |
| 9 | Shortness Of Breath | 52,220 | 2.9% | Vomiting | 13,541 | 3.4% |
| 10 | Vomiting | 50,461 | 2.8% | Altered Mental Status | 12,192 | 3.0% |

**Supplementary Table S2:**
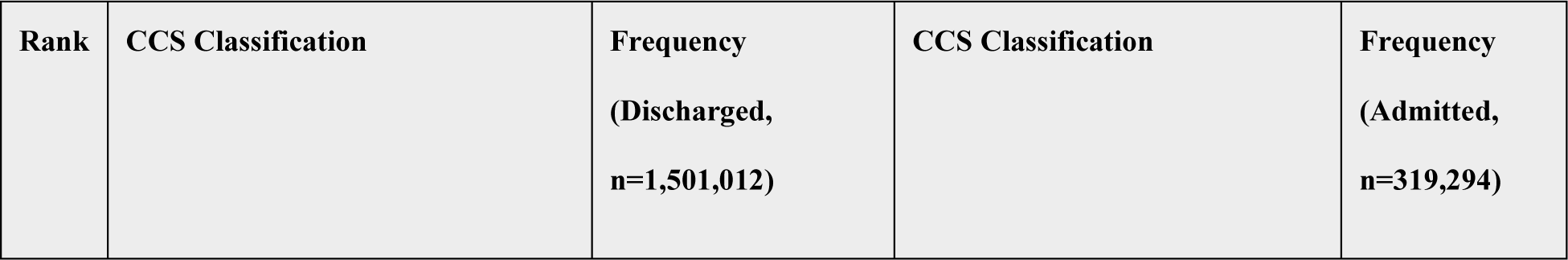
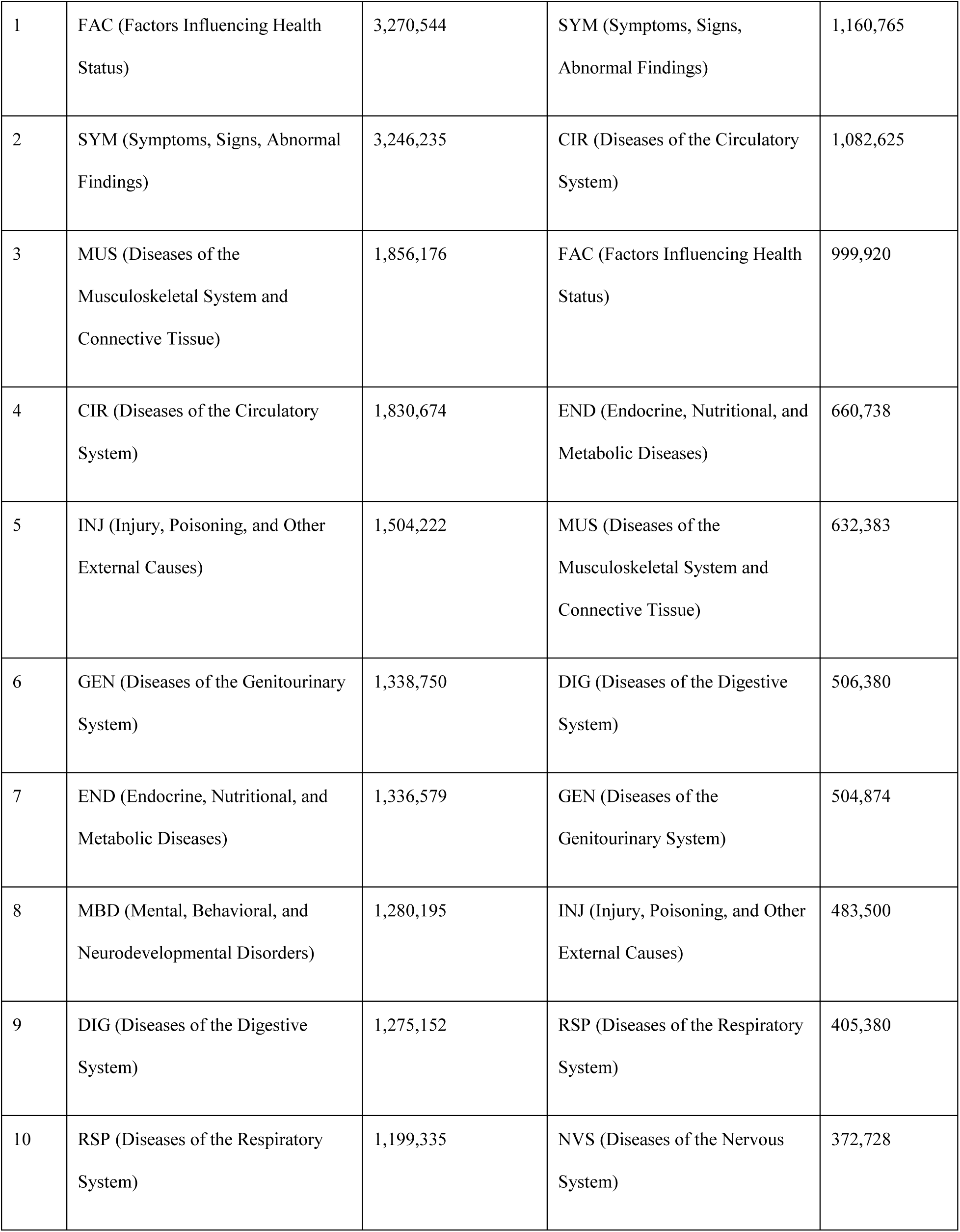
most common past medical history CCS categories among discharged and admitted patients.

| Rank | CCS Classification | Frequency<br>(Discharged,<br>n=1,501,012) | CCS Classification | Frequency<br>(Admitted,<br>n=319,294) |
| --- | --- | --- | --- | --- |
| 1 | FAC (Factors Influencing Health Status) | 3,270,544 | SYM (Symptoms, Signs, Abnormal Findings) | 1,160,765 |
| 2 | SYM (Symptoms, Signs, Abnormal Findings) | 3,246,235 | CIR (Diseases of the Circulatory System) | 1,082,625 |
| 3 | MUS (Diseases of the Musculoskeletal System and Connective Tissue) | 1,856,176 | FAC (Factors Influencing Health Status) | 999,920 |
| 4 | CIR (Diseases of the Circulatory System) | 1,830,674 | END (Endocrine, Nutritional, and Metabolic Diseases) | 660,738 |
| 5 | INJ (Injury, Poisoning, and Other External Causes) | 1,504,222 | MUS (Diseases of the Musculoskeletal System and Connective Tissue) | 632,383 |
| 6 | GEN (Diseases of the Genitourinary System) | 1,338,750 | DIG (Diseases of the Digestive System) | 506,380 |
| 7 | END (Endocrine, Nutritional, and Metabolic Diseases) | 1,336,579 | GEN (Diseases of the Genitourinary System) | 504,874 |
| 8 | MBD (Mental, Behavioral, and Neurodevelopmental Disorders) | 1,280,195 | INJ (Injury, Poisoning, and Other External Causes) | 483,500 |
| 9 | DIG (Diseases of the Digestive System) | 1,275,152 | RSP (Diseases of the Respiratory System) | 405,380 |
| 10 | RSP (Diseases of the Respiratory System) | 1,199,335 | NVS (Diseases of the Nervous System) | 372,728 |

## Machine Learning Models Hyper-Parameters Tuning

### XGBoost Model Performance and Hyperparameter Tuning

Hyperparameter tuning for the XGBoost model showed that admission prediction performance, measured by AUC, improved with specific configurations (**Supplementary Table S3**). The highest AUC score of 0.871 was achieved with n_estimators=1000, learning_rate=0.1, and max_depth=6, indicating this as the optimal setting. Performance generally increased with greater n_estimators and max_depth, particularly when paired with a moderate learning rate, reflecting the model’s sensitivity to deeper trees and a balanced learning pace.

**Supplementary Table S3:**
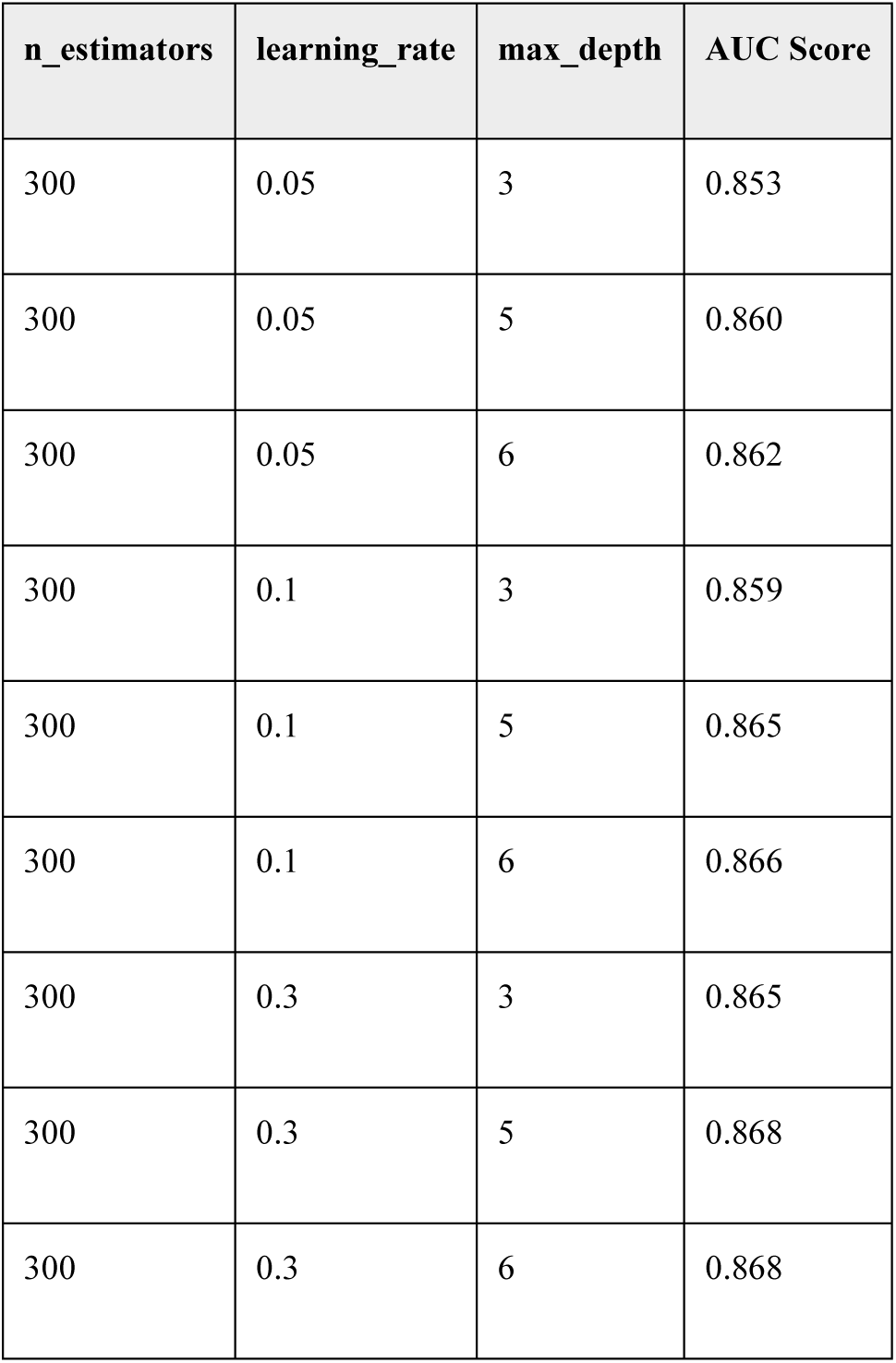
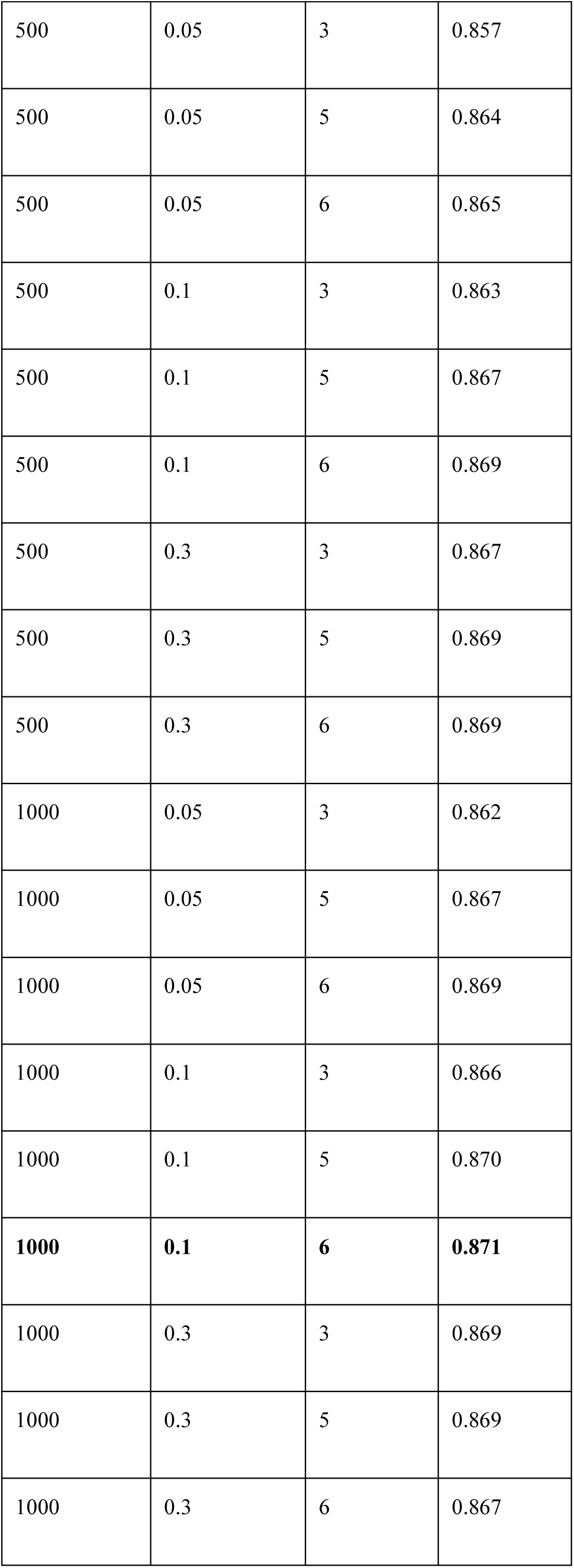
The table displays the AUC scores for XGBoost model performance on test data, evaluated across different hyperparameter settings. Each row represents a unique combination of n_estimators, learning_rate, and max_depth.

### Single Feature Analysis

Single feature analysis showed that “CCS set” (paste medical history), “Age,” and “ESI” had the top three highest predictive power, with AUC scores of 0.773, 0.726, and 0.707, respectively. In contrast, features like “Sex” and “Respirations” showed minimal predictive value, emphasizing the varying strength of individual features in admission prediction (**Supplementary Figure S1**).

**Supplementary Figure S1:**
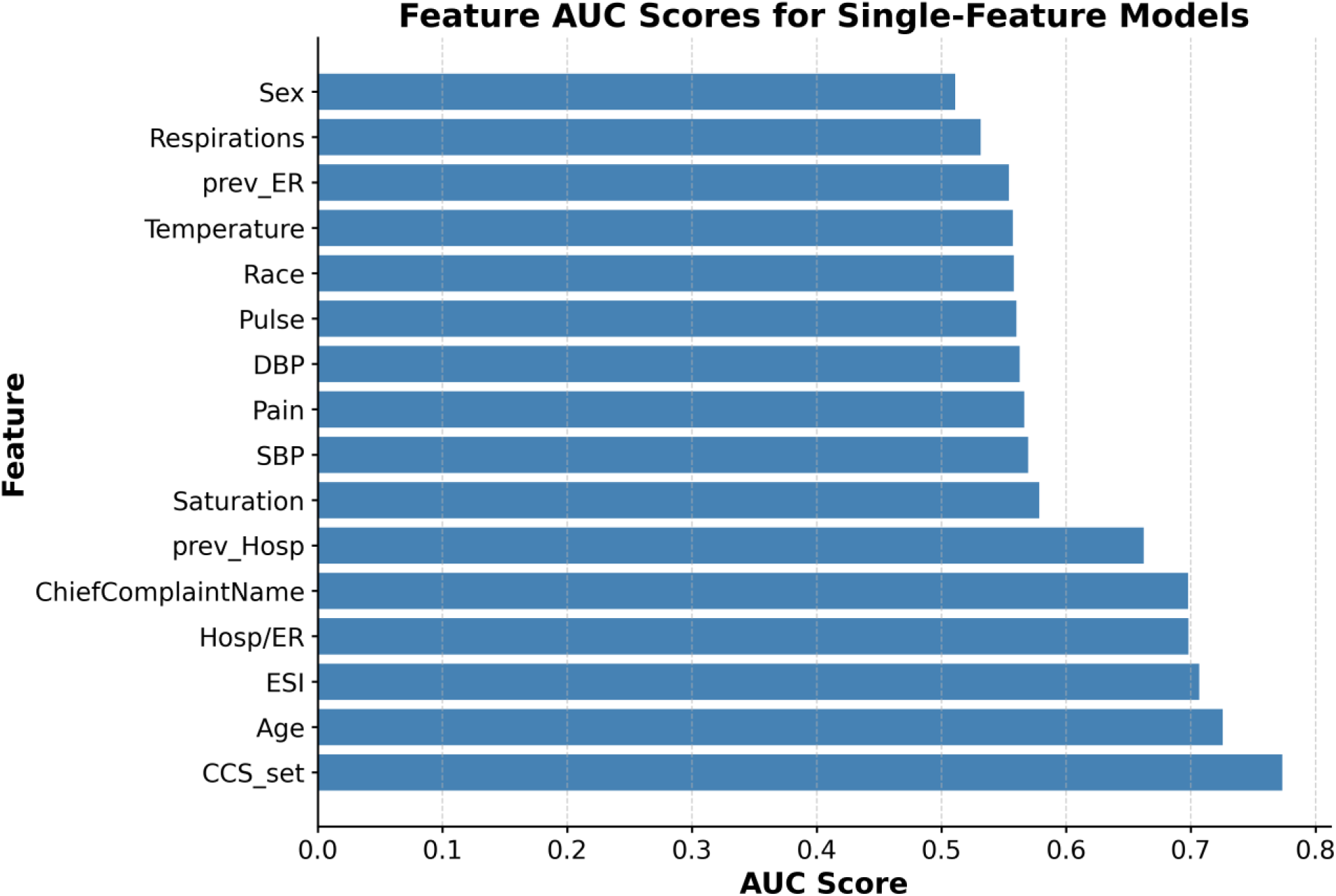
This figure displays the AUC scores for individual features in the XGBoost model, highlighting each feature’s standalone predictive power for admission.

### Hyperparameter Tuning for Bio-clinical-BERT: Initial Exploration

We explored Bio-clinical-BERT performance with different learning rates (lr), batch sizes (bs), and weight decay (wd) using a random set of 10k samples from the training set and a random set of 2k samples from the internal validation set. Performance was evaluated using AUC. The highest AUC achieved was 0.819 with a learning rate of 10^-5^, batch size of 8, and weight decay of 0.01 (**Supplementary Table S4**).

**Supplementary Table S4:** Bio-clinical-BERT hyperparameter tuning results.

| Learning Rate (lr) | Batch Size (bs) | Weight Decay (wd) | AUC |
| --- | --- | --- | --- |
| 0.00001 | 8 | 0.01 | 0.819 |
| 0.00005 | 128 | 0.01 | 0.819 |
| 0.00005 | 64 | 0.01 | 0.818 |
| 0.00002 | 16 | 0.10 | 0.818 |
| 0.00002 | 32 | 0.01 | 0.818 |
| 0.00001 | 16 | 0.01 | 0.817 |
| 0.00005 | 128 | 0.10 | 0.817 |
| 0.00001 | 8 | 0.10 | 0.816 |
| 0.00002 | 8 | 0.10 | 0.816 |
| 0.00002 | 16 | 0.01 | 0.816 |
| 0.00002 | 64 | 0.10 | 0.815 |
| 0.00002 | 8 | 0.01 | 0.815 |
| 0.00005 | 16 | 0.01 | 0.815 |
| 0.00005 | 32 | 0.10 | 0.813 |
| 0.00001 | 32 | 0.10 | 0.812 |
| 0.00005 | 16 | 0.10 | 0.812 |
| 0.00001 | 16 | 0.10 | 0.811 |
| 0.00002 | 64 | 0.01 | 0.810 |
| 0.00001 | 32 | 0.01 | 0.809 |
| 0.00005 | 32 | 0.01 | 0.809 |
| 0.00005 | 8 | 0.10 | 0.806 |
| 0.00005 | 64 | 0.10 | 0.806 |
| 0.00005 | 256 | 0.01 | 0.801 |
| 0.00005 | 256 | 0.10 | 0.800 |
| 0.00002 | 128 | 0.10 | 0.800 |
| 0.00001 | 64 | 0.01 | 0.800 |
| 0.00005 | 8 | 0.01 | 0.799 |
| 0.00001 | 64 | 0.10 | 0.793 |
| 0.00002 | 128 | 0.01 | 0.792 |
| 0.00002 | 256 | 0.01 | 0.777 |
| 0.00002 | 256 | 0.10 | 0.772 |
| 0.00001 | 128 | 0.01 | 0.753 |
| 0.00001 | 128 | 0.10 | 0.746 |
| 0.00001 | 256 | 0.01 | 0.706 |
| 0.00001 | 256 | 0.10 | 0.697 |
| 0.00005 | 32 | 0.10 | 0.478 |

The model was further evaluated with an expanded dataset of 50k training samples and 10k test samples. The highest AUC achieved was 0.840 with a learning rate 10^-5^, batch size of 8, and weight decay of 0.01 (**Supplementary Table S5**).

Given the operational constraints and efficiency considerations, we selected the top-performing configuration with a batch size of 256 for the final model. This configuration achieved an AUC of 0.838 with a learning rate of 5×10^-6^ and weight decay of 0.10, balancing strong performance with computational scalability.

**Supplementary Table S5:**
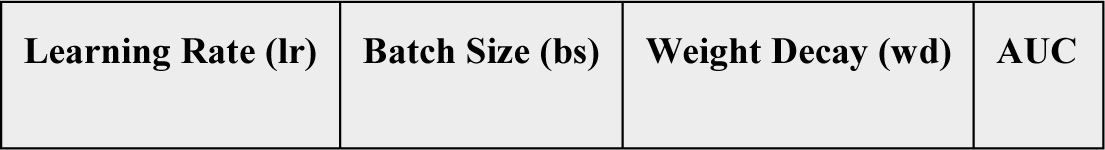
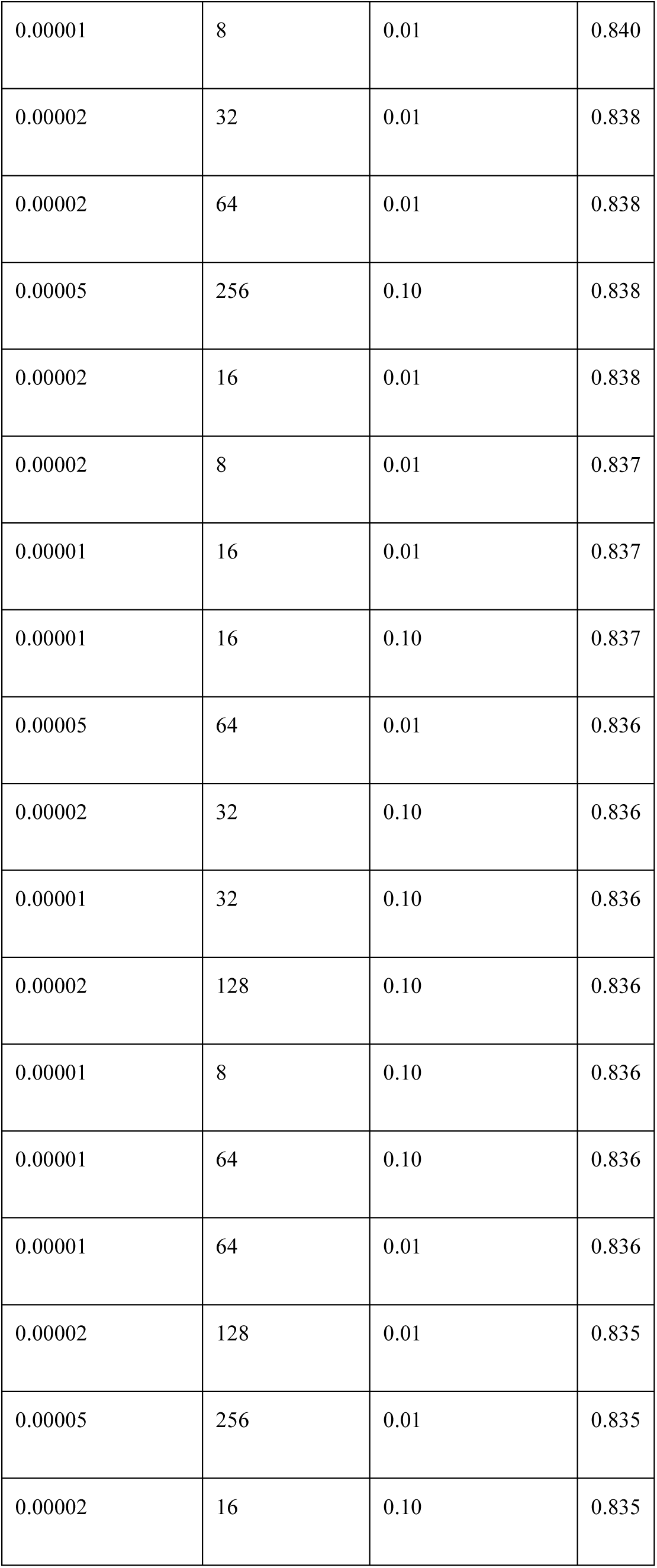
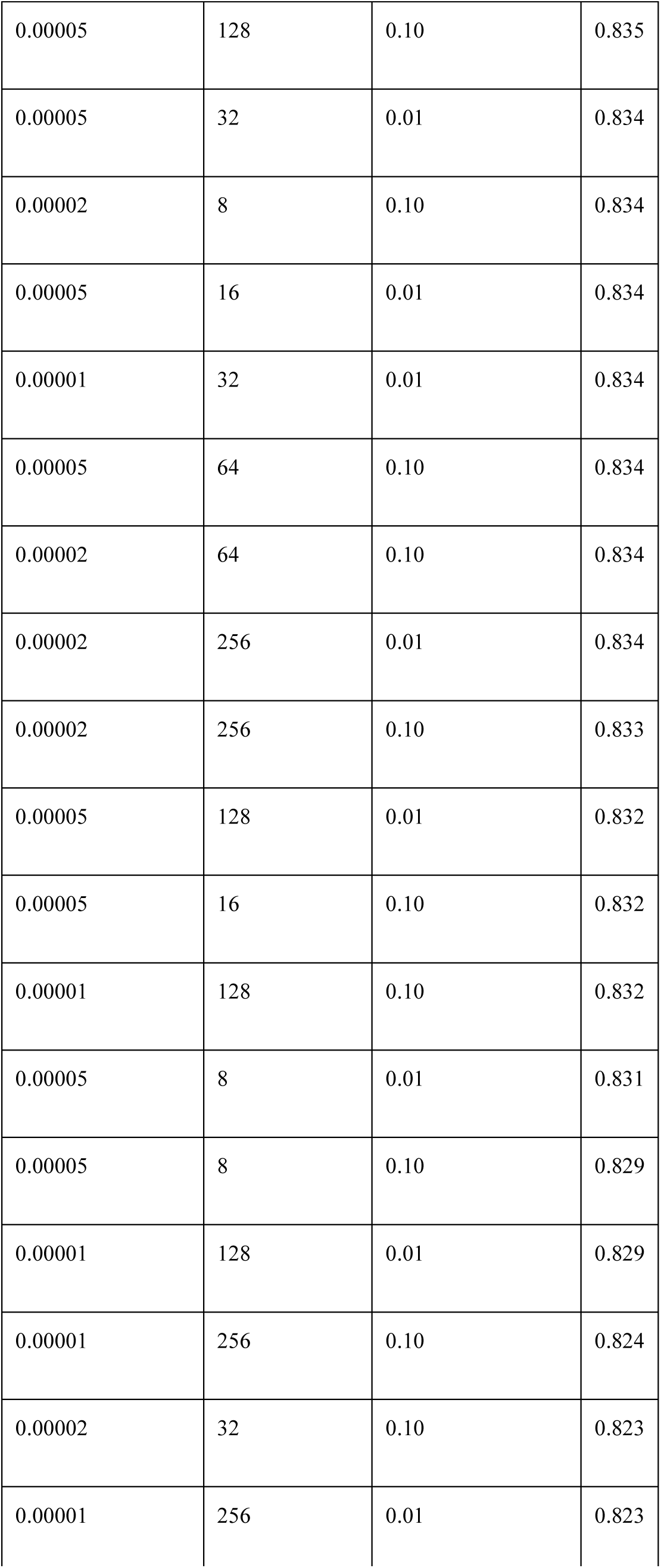
Hyperparameter tuning results (training set of 50,000 visits).

| Learning Rate (lr) | Batch Size (bs) | Weight Decay (wd) | AUC |
| --- | --- | --- | --- |
| 0.00001 | 8 | 0.01 | 0.840 |
| 0.00002 | 32 | 0.01 | 0.838 |
| 0.00002 | 64 | 0.01 | 0.838 |
| 0.00005 | 256 | 0.10 | 0.838 |
| 0.00002 | 16 | 0.01 | 0.838 |
| 0.00002 | 8 | 0.01 | 0.837 |
| 0.00001 | 16 | 0.01 | 0.837 |
| 0.00001 | 16 | 0.10 | 0.837 |
| 0.00005 | 64 | 0.01 | 0.836 |
| 0.00002 | 32 | 0.10 | 0.836 |
| 0.00001 | 32 | 0.10 | 0.836 |
| 0.00002 | 128 | 0.10 | 0.836 |
| 0.00001 | 8 | 0.10 | 0.836 |
| 0.00001 | 64 | 0.10 | 0.836 |
| 0.00001 | 64 | 0.01 | 0.836 |
| 0.00002 | 128 | 0.01 | 0.835 |
| 0.00005 | 256 | 0.01 | 0.835 |
| 0.00002 | 16 | 0.10 | 0.835 |
| 0.00005 | 128 | 0.10 | 0.835 |
| 0.00005 | 32 | 0.01 | 0.834 |
| 0.00002 | 8 | 0.10 | 0.834 |
| 0.00005 | 16 | 0.01 | 0.834 |
| 0.00001 | 32 | 0.01 | 0.834 |
| 0.00005 | 64 | 0.10 | 0.834 |
| 0.00002 | 64 | 0.10 | 0.834 |
| 0.00002 | 256 | 0.01 | 0.834 |
| 0.00002 | 256 | 0.10 | 0.833 |
| 0.00005 | 128 | 0.01 | 0.832 |
| 0.00005 | 16 | 0.10 | 0.832 |
| 0.00001 | 128 | 0.10 | 0.832 |
| 0.00005 | 8 | 0.01 | 0.831 |
| 0.00005 | 8 | 0.10 | 0.829 |
| 0.00001 | 128 | 0.01 | 0.829 |
| 0.00001 | 256 | 0.10 | 0.824 |
| 0.00002 | 32 | 0.10 | 0.823 |
| 0.00001 | 256 | 0.01 | 0.823 |

**Supplementary Table S6:** performance metrics of the XGBoost model at different probability thresholds. Metrics include F1, accuracy, sensitivity, specificity, and predictive values with 95% confidence intervals.

| Threshold | Accuracy | F1 Score | Sensitivity | Specificity | NPV | PPV |
| --- | --- | --- | --- | --- | --- | --- |
| 0.10 | 69.4% (69.0% - 69.9%) | 0.53 (0.52 - 0.54) | 90.0% (89.4% - 90.6%) | 64.5% (64.0% - 65.0%) | 96.4% (96.2% - 96.7%) | 37.7% (37.1% - 38.4%) |
| 0.19<br>(Youden's index) | 79.7% (79.4% - 80.1%) | 0.60 (0.59 - 0.61) | 78.9% (78.1% - 79.8%) | 79.9% (79.5% - 80.4%) | 94.1% (93.8% - 94.3%) | 48.4% (47.7% - 49.2%) |
| 0.30 | 84.2% (83.9% - 84.6%) | 0.62 (0.61 - 0.63) | 66.8% (65.9% - 67.8%) | 88.4% (88.1% - 88.7%) | 91.8% (91.5% - 92.1%) | 57.9% (56.9% - 58.8%) |
| 0.50 | 85.9% (85.5% - 86.2%) | 0.55 (0.54 - 0.56) | 45.3% (44.3% - 46.4%) | 95.6% (95.4% - 95.8%) | 88.0% (87.6% - 88.3%) | 70.9% (69.7% - 72.0%) |
| 0.70 | 84.3% (84.0% - 84.7%) | 0.37 (0.36 - 0.38) | 23.6% (22.8% - 24.5%) | 98.8% (98.7% - 98.9%) | 84.4% (84.1% - 84.8%) | 82.6% (81.0% - 84.1%) |
| 0.90 | 81.7% (81.4% - 82.1%) | 0.10 (0.10 - 0.11) | 5.5% (5.0% - 6.0%) | 99.9% (99.9% - 99.9%) | 81.6% (81.2% - 81.9%) | 94.1% (91.9% - 96.0%) |

**Supplementary Table S7:** Performance metrics of the NLP model at different probability thresholds. Metrics include F1, accuracy, sensitivity, specificity, and predictive values with 95% confidence intervals.

| Threshold | Accuracy | F1 Score | Sensitivity | Specificity | NPV | PPV |
| --- | --- | --- | --- | --- | --- | --- |
| 0.10 | 64.2% (63.7% - 64.6%) | 0.50 (0.49 - 0.50) | 90.7% (90.1% - 91.3%) | 57.7% (57.2% - 58.2%) | 96.3% (96.0% - 96.5%) | 34.1% (33.5% - 34.7%) |
| 0.19 (Youden's index) | 76.4% (76.1% - 76.8%) | 0.57 (0.56 - 0.58) | 79.7% (78.8% - 80.5%) | 75.7% (75.2% - 76.1%) | 93.9% (93.6% - 94.2%) | 44.2% (43.4% - 44.9%) |
| 0.30 | 83.1% (82.8% - 83.4%) | 0.60 (0.59 - 0.61) | 65.4% (64.4% - 66.3%) | 87.4% (87.1% - 87.7%) | 91.3% (91.0% - 91.6%) | 55.6% (54.6% - 56.5%) |
| 0.50 | 85.6% (85.3% - 85.9%) | 0.55 (0.54 - 0.56) | 45.1% (44.1% - 46.2%) | 95.4% (95.2% - 95.6%) | 87.8% (87.5% - 88.1%) | 70.2% (69.0% - 71.4%) |
| 0.70 | 84.6% (84.3% - 84.9%) | 0.40 (0.39 - 0.41) | 26.7% (25.8% - 27.6%) | 98.6% (98.5% - 98.7%) | 84.8% (84.5% - 85.1%) | 82.3% (80.9% - 83.7%) |
| 0.90 | 82.4% (82.1% - 82.8%) | 0.19 (0.17 - 0.20) | 10.3% (9.6% - 10.9%) | 99.8% (99.8% - 99.9%) | 82.2% (81.8% - 82.5%) | 93.0% (91.4% - 94.5%) |

